# Digging deeper into GWAS signal using GRIN implicates additional genes contributing to suicidal behavior

**DOI:** 10.1101/2022.04.20.22273895

**Authors:** Kyle A. Sullivan, Matthew Lane, Mikaela Cashman, J. Izaak Miller, Mirko Pavicic, Angelica M. Walker, Ashley Cliff, Jonathon Romero, Xuejun Qin, Jennifer Lindquist, Niamh Mullins, Anna Docherty, Hilary Coon, Douglas M. Ruderfer, International Suicide Genetics Consortium, VA Million Veteran Program, MVP Suicide Exemplar Workgroup, Michael R. Garvin, John P. Pestian, Allison E. Ashley-Koch, Jean C. Beckham, Benjamin McMahon, David W. Oslin, Nathan A. Kimbrel, Daniel A. Jacobson, David Kainer

**Affiliations:** Computational and Predictive Biology, Oak Ridge National Laboratory, Oak Ridge, TN, USA; The Bredesen Center for Interdisciplinary Research and Graduate Education, University of Tennessee Knoxville, Knoxville, TN, USA; Durham Veterans Affairs Health Care System, Durham, NC, USA; Duke University School of Medicine, Duke University, Durham, NC, USA; Department of Genetics and Genomic Sciences, Icahn School of Medicine at Mount Sinai, New York City, NY, USA; Department of Psychiatry, Icahn School of Medicine at Mount Sinai, New York City, NY, USA; Department of Psychiatry, University of Utah School of Medicine, Salt Lake City, UT, USA; Huntsman Mental Health Institute, University of Utah School of Medicine, Salt Lake City, UT, USA; Division of Genetic Medicine, Vanderbilt University Medical Center, Nashville, TN, USA; Vanderbilt Genetics Institute, Vanderbilt University Medical Center, Nashville, TN, USA; Department of Biomedical Informatics, Vanderbilt University Medical Center, Nashville, TN, USA; Department of Psychiatry and Behavioral Sciences, Vanderbilt University Medical Center, Nashville, TN, USA; Cincinnati Children’s Hospital Medical Center, University of Cincinnati, Cincinnati, OH, USA; Department of Psychiatry and Behavioral Sciences, Duke University Medical Center, Durham, NC, USA; VISN 6 Mid-Atlantic Mental Illness Research, Durham Veterans Affairs Health Care System, Durham, NC, USA; Theoretical Biology and Biophysics, Los Alamos National Laboratory, Los Alamos, NM, USA; VISN 4 Mental Illness Research, Education, and Clinical Center, Center of Excellence, Corporal Michael J. Crescenz VA Medical Center, Philadelphia, PA, USA; Department of Psychiatry, Perelman School of Medicine, University of Pennsylvania, PA, USA; VA Health Services Research and Development Center of Innovation to Accelerate Discovery and Practice Transformation, Durham, NC

## Abstract

Genome-wide association studies (GWAS) identify genetic variants underlying complex traits but are limited by stringent genome-wide significance thresholds. Here we dramatically relax GWAS stringency by orders of magnitude and apply GRIN (Gene set Refinement through Interacting Networks), which increases confidence in the expanded gene set by retaining genes strongly connected by biological networks from diverse lines of evidence. From multiple GWAS summary statistics of suicide attempt, a complex psychiatric phenotype, GRIN identified additional genes that replicated across independent cohorts and retained genes that were more biologically interrelated despite a relaxed significance threshold. We present a conceptual model of how these retained genes interact through neurobiological pathways to influence suicidal behavior and identify existing drugs associated with these pathways that would not have been identified under traditional GWAS thresholds. We demonstrate that GRIN is a useful community resource for improving the signal to noise ratio of GWAS results.

## Introduction

Genome-wide association studies (GWAS) can identify the genetic basis of complex phenotypes but interpreting the results of these studies is often challenging. SNP-level results from GWAS often need gene assignment for understanding downstream functions, but SNP-to-gene assignment is challenging for intergenic SNPs^1–4^. Moreover, while the resulting gene set may contain genes that affect the phenotype, these “true positive” genes are often interspersed with false positives as a consequence of confounding experimental factors and incorrect SNP-to-gene assignments. False positive genes can mislead efforts to understand the genetic architecture of a trait, so it is important that they are filtered out. This becomes even more crucial for GWAS that produce few associated genes at traditional genome-wide significance (i.e., *p* < 5e^-8^). For these cases it is often necessary to investigate SNPs at less stringent *p*-value thresholds to find relevant loci, at the risk of introducing false positives.

Polygenic traits are typically influenced by genes from multiple pathways working in concert with each other within the broader biological system^5^. Given enough information from diverse experimental sources, it should be possible to find functional lines of biological evidence connecting any pair of truly causal genes to each other more strongly than pairs of random genes. When considered as a set (*e*.*g*., from a GWAS or differential gene expression), the true positive causal genes are very likely to be functionally connected to one another. Conversely, false positive genes from a GWAS will likely be random and therefore far less functionally connected to the other genes in the set. Given a gene set, one could conceivably determine which genes are most likely to be false positives if the gene set can be parsed effectively by including thorough gene interaction information.

To solve this important biological problem, we present GRIN – Gene set Refinement through Interacting Networks – an approach that removes biologically disparate, false positive genes in a gene set based upon biological network topology. GRIN uses a network representation of system-wide gene-to-gene interactions from diverse experimental sources. Starting from a user-defined gene set, GRIN explores the network topology to determine how strongly these genes are interconnected. Next, GRIN compares the connectivity among these genes to the connectivity found among random genes to determine which of the user’s genes are likely to be false positives. We validated GRIN by testing its ability to separate well-characterized, functionally related “gold standard” genes from random genes using a large multiplex network^6^.

We then applied GRIN to suicide attempt GWAS summary statistics, a complex psychiatric phenotype. Even though heritability estimates range from 17-55%^7,8^, this psychiatric disorder has elicited few genome-wide significant variants to date^8–11^. We analyzed independent suicide attempt GWAS results from the Million Veteran Program (MVP^11^) and the International Suicide Genetics Consortium (ISGC^9^). As few variants were significant at traditional genome-wide significance, we explored SNPs at a less stringent threshold and used multiple SNP-to-gene assignment methods to elucidate the underlying mechanisms of the heritable components of suicidality. GRIN identified genes with higher probability of contributing to suicide attempt pathophysiology, reducing false positive genes. Thus, we demonstrate GRIN’s utility in identifying key biological mechanisms from GWAS signals, and subsequently identify putative drug targets for future suicide prevention studies.

## Results

### GRIN workflow

A summary of the GRIN workflow is presented in **Figure 1**. GRIN inputs include a gene set and a previously generated multiplex network. In Stage 1, all experimentally-derived (*e*.*g*., GWAS) “seed” genes are ranked based upon biological network connectivity. GRIN also performs this task on random gene sets of equivalent size to produce an empirical null rank distribution. In Stage 2, a sliding window is used to compare the ordered ranks of experimentally-derived genes to the equivalent ordered ranks within the null distribution using the Mann-Whitney U test, and the *p*-value of the Mann-Whitney U test is plotted for each window to form a curve. The elbow of this curve indicates the cutoff point at which the ranks are equivalent between the seed gene set and the null distribution (indicating low functional interrelatedness), and genes following this cutoff point are filtered out.

**Figure 1.**
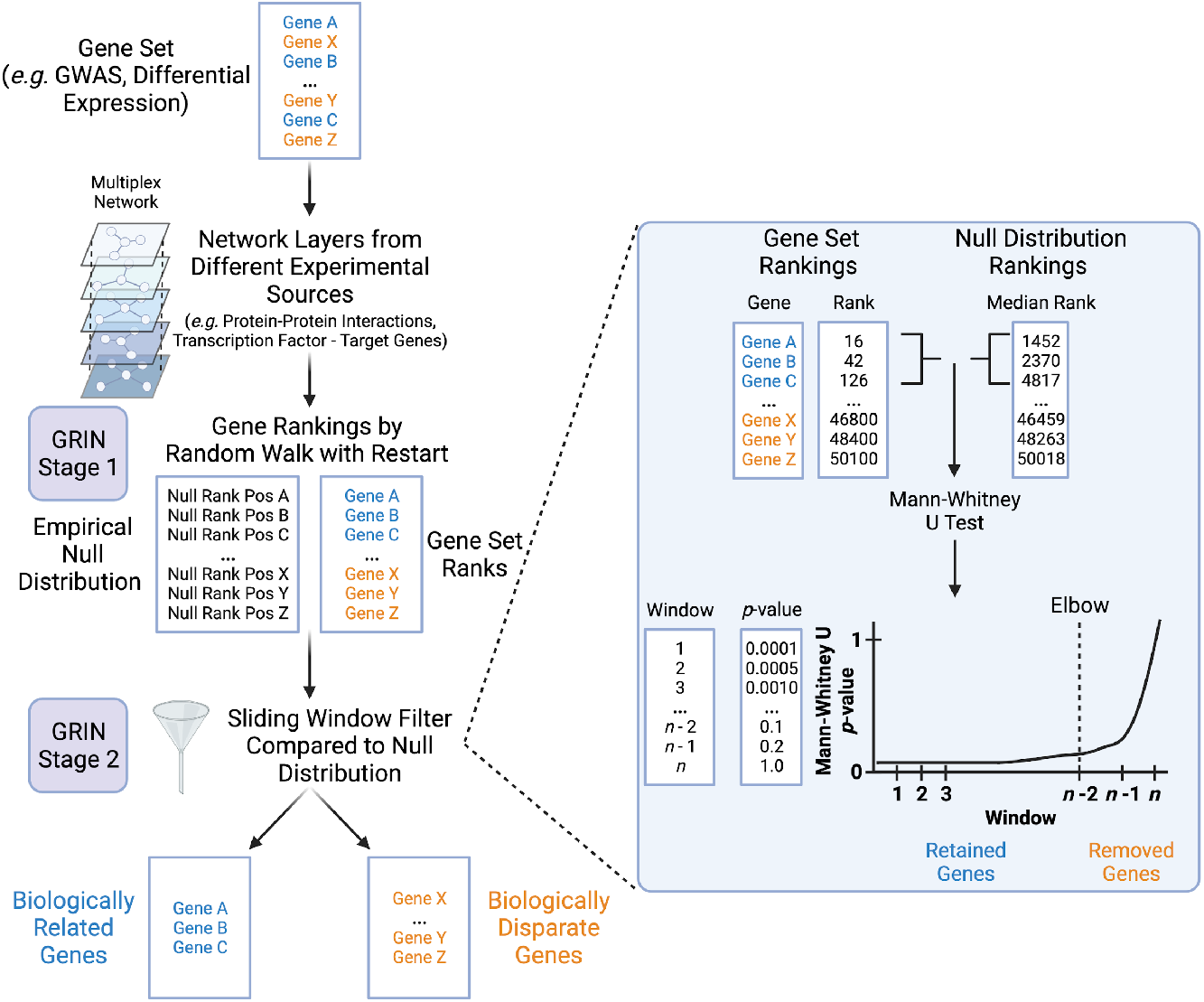
Overview of GRIN – Gene set Refinement through Interacting Networks. In stage 1, GRIN requires a network composed of gene-gene connections, preferably assembled from multiple experimental data sources. Next, the network is explored starting from all genes in the user’s gene set using the random walk with restart algorithm, resulting in a rank-ordered list of all genes in the network based on the frequency in which they were visited. The ordered ranks of the user’s genes are used for stage 2, where they are compared to ordered ranks obtained from running Stage 1 on 100 random gene sets of the same size (empirical null distribution). Using the Mann-Whitney U test, a window of the user’s gene set ranks is compared to the empirical null distribution of ranks to determine if the user’s gene rankings come from the same distribution as random genes’ rankings. After using a sliding window to compare the gene ranks in this manner, GRIN identifies a set of biologically related genes at the point at which the ranks of the user’s gene list begin to approximate the ranks of the null distribution (elbow point of the curve). Genes prior to the sliding window at the elbow point are retained as their ranks deviate from the null distribution, while genes beyond this point are removed.

### GRIN accurately removes false positive genes as determined by benchmarking with biologically interrelated, “gold standard” gene sets

We first tested GRIN on 30 biologically-interrelated, “gold standard” gene sets mixed with an equivalent amount of randomly drawn genes (1:1 ratio), and repeated this 100 times per gold standard gene set (**Fig. 2A, Supplementary Table 1**). GRIN Stage 1 ranked gold standard genes more highly than most random genes in any given test set, achieving median area under the receiver operating characteristic curve (AUROC) and median area under the precision-recall curve (AUPRC) values of 0.950 ± 0.059 and 0.896 ± 0.084 respectively across the 30 gold standard gene sets (**Fig. 2B-C**). This indicated highly accurate gene ranking in GRIN Stage 1, which is necessary for successful outcomes at Stage 2. GRIN Stage 2 effectively classified true and false positive genes at the cutoff point (median precision 0.810 ± 0.134; recall 0.914 ± 0.167; specificity 0.880 ± 0.197; **Fig. 2D, Supplementary Fig. 1**). On average, 46.0% of the gene set was discarded as noise (0.460 ± 0.143), and less than 9% of true positive gold genes were discarded (median recall 0.914).

**Figure 2.**
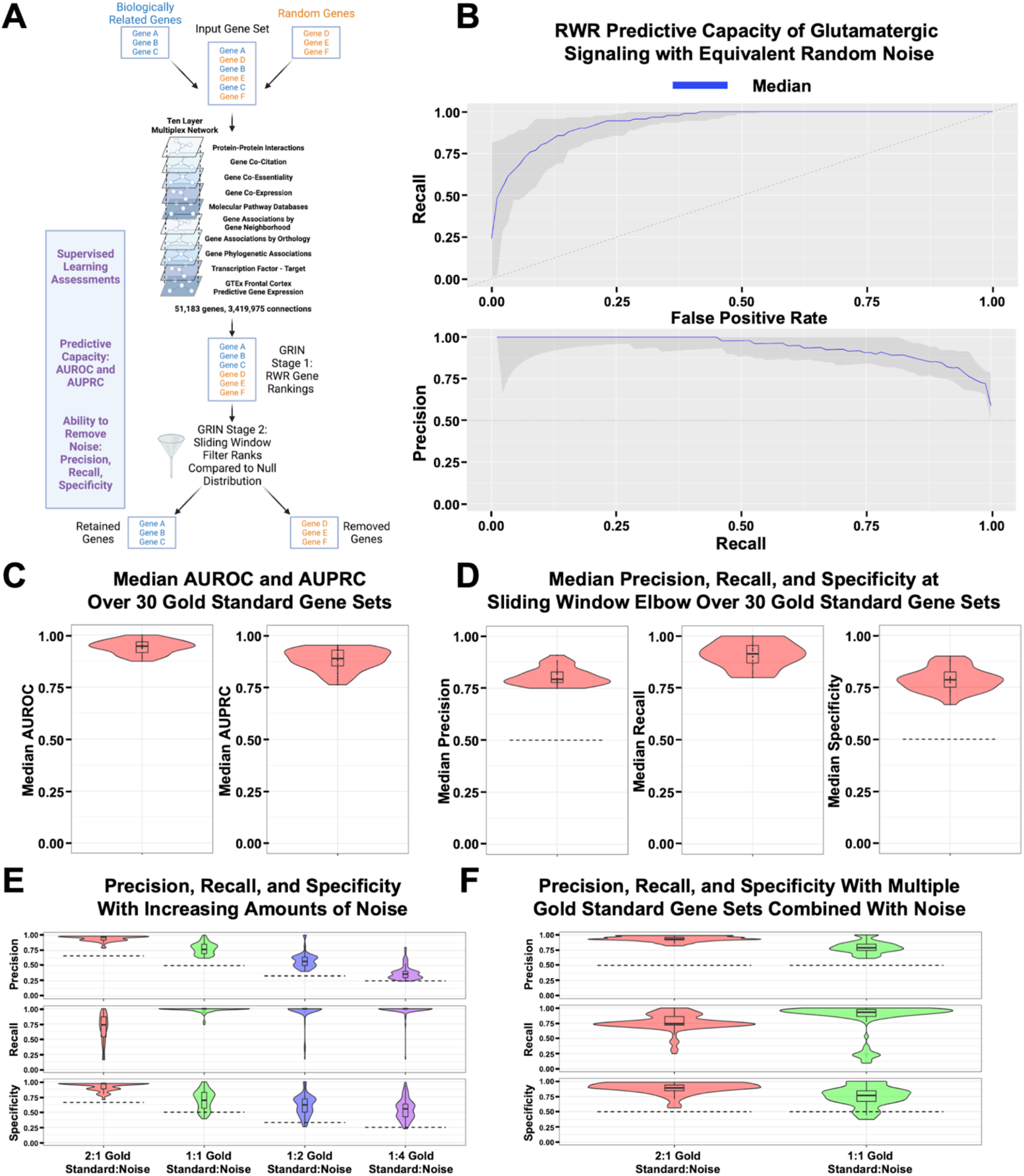
GRIN retains biologically interrelated genes. **A**. Workflow for assessing the capacity of GRIN to identify biologically related genes from well-defined, “gold standard” gene sets (GO, OMIM, DisGeNET) mixed with random genes. Figure made with BioRender.com. **B**. Receiver-operating characteristic (ROC) curve and precision-recall curves (PRC) for GRIN. The blue line represents the mean of GRIN performance over 100 sets of glutamatergic signaling genes (GO:0035249) intermixed with an equivalent number of random genes from the multiplex network. Gray values indicate maximum and minimum values over 100 sets, and dotted lines indicate classification values based on random chance. **C**. Median area under ROC (AUROC) and PRC (AUPRC) values for 100 sets of random genes intermixed with a gold standard gene set for 30 distinct gold standard gene sets. **D**. Median precision, recall, and specificity values for 100 sets of a gold standard gene set intermixed with an equivalent number of random genes, repeated for 30 distinct gold standard gene sets. **E**. Violin and box plots of precision, recall, and specificity values for 100 sets of dopaminergic synaptic signaling (GO:0001963) gold standard genes intermixed with different ratios of random genes (Noise). Median precision: 0.970 (2:1), 0.767 (1:1), 0.568 (1:2), 0.360 (1:4). Median recall: 0.735 (2:1), 1.00 (1:1), 1.00 (1:2), 1.00 (1:4). Median specificity: 0.970 (2:1), 0.696 (1:1), 0.620 (1:2), 0.554 (1:4) **F**. Precision, recall, and specificity values for 100 sets of dopaminergic synaptic signaling (GO:0001963) and myelination (GO:0022010) gold standard genes mixed with a ratio of 50% noise (2:1 Gold Std:Noise) or an equivalent ratio (1:1 Gold Std:Noise) of random genes.

Next, we tested GRIN’s ability to refine gene sets containing varying proportions of random genes. When gold standard genes outnumbered random genes by 2:1, precision (0.967 ± 0.060), recall (0.735 ± 0.331), and specificity (0.971± 0.088) were consistently high compared to expected values from random classification (precision = 0.66, specificity = 0.66). Precision, recall, and specificity values decreased as the ratio of gold genes to random genes in the gene sets decreased (1:1, 1:2, 1:4) but were consistently better than random chance (**Fig. 2E, Supplementary Fig. 2**).

Finally, we tested whether GRIN could retain multiple distinct functional groups when mixed with random genes. This test case is important since a real-world gene set for a complex trait is likely to contain multiple functional groups. When given gene sets with two distinctive biological functions (“dopaminergic synaptic signaling” and “myelination”) at 1:1 or 2:1 ratios of gold standard genes to random genes, GRIN generated much better precision and specificity compared to random chance while retaining high values for recall (2:1 merged gold standard:random – precision: 0.931 ± 0.042, recall: 0.742 ± 0.141, specificity: 0.891 ± 0.094; 1:1 merged gold standard:random – precision: 0.789 ± 0.103, recall: 0.930 ± 0.116, specificity: 0.767 ± 0.174; **Fig. 2F**).

### GRIN retains biologically interrelated genes from the Million Veteran Program (MVP) suicide attempt GWAS at less stringent significance thresholds

After benchmarking GRIN on gold standard gene sets, we applied it to suicide attempt GWAS summary statistics from the Million Veteran Program (MVP^11^) to understand which genes contribute to psychiatric pathophysiology in this military veteran population. We utilized both conventional MAGMA^4^ and H-MAGMA^1^ solely for gene assignment from SNPs at different genome-wide significance thresholds (**Fig. 3A**). Only five SNPs were significant below the traditional threshold of genome-wide significance (*p* < 5e^-8^) which were assigned to three total genes (*SPATA17, TSHZ2*, and *ENSG00000227705*; **Supplementary Tables 2 and 3**). In order to explore additional genes contributing to suicide attempt pathophysiology, we explored markedly less stringent *p*-value thresholds which expanded the GWAS-derived gene sets. We only examined SNPs and resulting genes at the *p* < 1e^-5^ threshold since the numbers of genes obtained at less stringent thresholds became unwieldy to interpret in a unified context (**Supplementary Table 2**).

**Figure 3.**
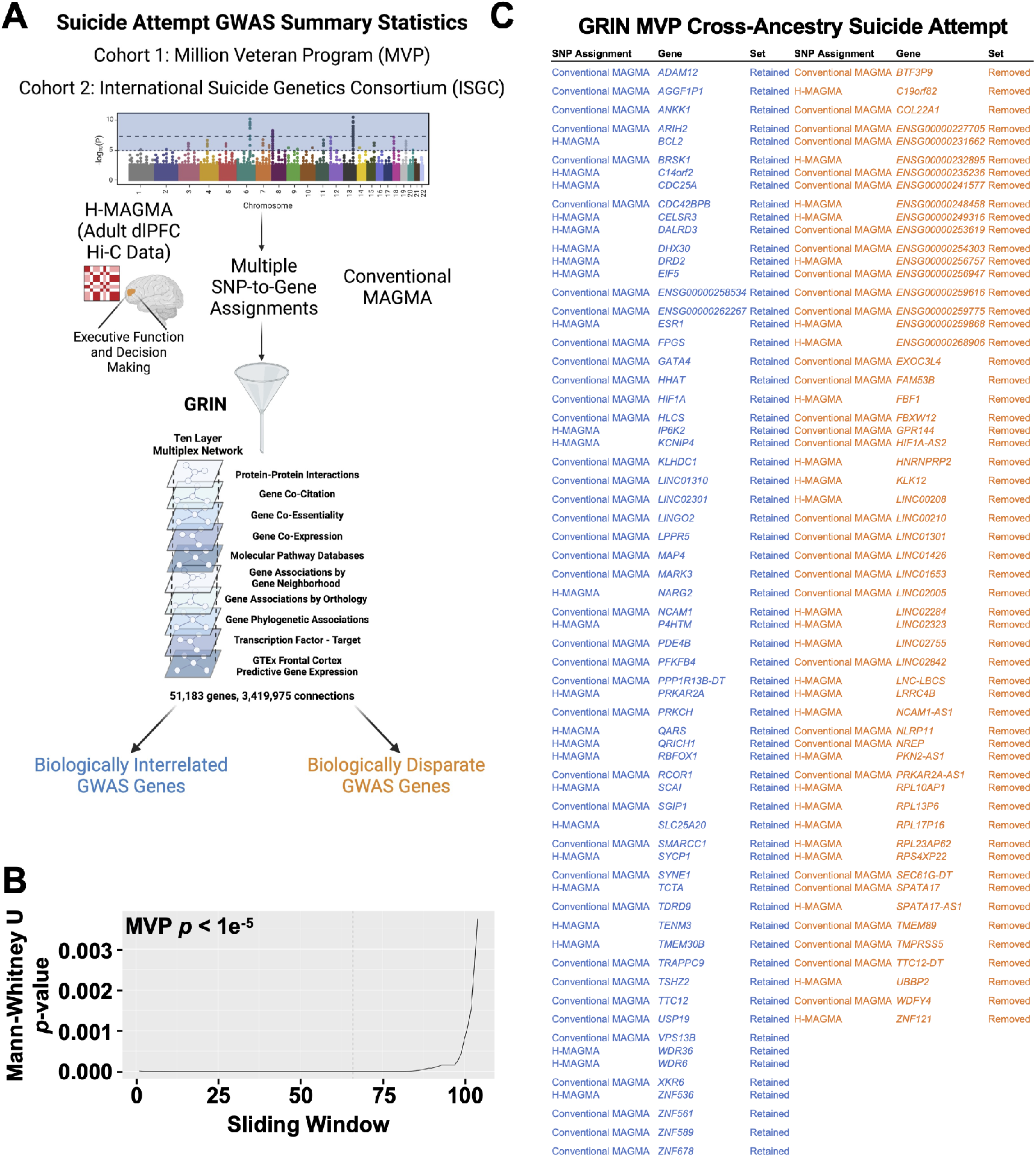
GRIN identifies biologically interrelated genes from suicide attempt summary statistics from United States veterans. **A**. Workflow for identifying biologically interrelated genes from genome-wide association study (GWAS) summary statistics of suicide attempt from Million Veteran Program (MVP) and International Suicide Genetics Consortium (ISGC) cohorts. Figure made with BioRender.com. **B**. Sliding window *p*-values of Mann-Whitney U test for the union of H-MAGMA and conventional MAGMA-assigned genes from MVP GWAS summary statistics using SNPs with a threshold of *p* < 1e^-5^. Dotted line indicates the elbow point of the curve used to determine which genes were retained by GRIN. **C**. Gene lists of retained (blue) and removed (orange) genes from MVP summary statistics as determined by GRIN based on H-MAGMA or conventional MAGMA SNP-to-gene assignment.

MVP SNPs were assigned to 122 genes at *p* < 1e^-5^ (**Supplementary Tables 2 and 4**). We applied GRIN to this gene set in order to limit the higher number of potential false positive genes introduced at this threshold. GRIN retained 65 genes and removed 57 genes from the gene set (**Fig. 3B-C, Supplementary Table 5**). Next, to determine if GRIN successfully retained biologically interrelated genes we used ToppGene^12^ to compare a variety of gene set enrichments (**Methods**) of MVP genes before and after GRIN. The 122 unfiltered genes were not significantly enriched for any GO terms but were enriched for 203 combined drug and disease enrichments, including “substance dependence” (DisGeNET C0038580; **Supplementary Table 6**). Intriguingly, after refining the gene set with GRIN, 1449 enrichments were significant from 65 retained genes, including 18 transcription factor binding sites and five GO molecular function enrichments (*e*.*g*., “adenyl ribonucleotide binding,” **Supplementary Table 7**). Conversely, GRIN removed 57 genes that manifested no significant enrichments. Thus, filtering MVP genes at a far less stringent GWAS significance threshold with GRIN resulted in a more functionally enriched gene set by improving the signal-to-noise ratio.

### GRIN retains biologically interrelated genes from multiple International Suicide Genetics Consortium (ISGC) GWAS at a less stringent significance threshold

We sought to identify which genes retained by GRIN from MVP were replicated in two independent, civilian suicide attempt GWAS compiled by the International Suicide Genetics Consortium (ISGC^9^). This study contained summary statistics from: 1) a European ancestry GWAS of suicide attempt in civilians (SA-EUR) and 2) a GWAS of the SA-EUR population conditioned by major depressive disorder (MDD) diagnosis status (SA-MDD). At a threshold of *p* < 5e^-8^, only seven SA-EUR genes and three SA-MDD genes were significantly associated (**Supplementary Tables 2-3**).

We then applied the same, less stringent significance threshold (*p* < 1e^-5^) to identify additional genes, and applied GRIN to remove potential false positives. Following SNP-to-gene assignment, 252 genes were identified from SA-EUR and 62 genes were identified from SA-MDD (**Supplementary Tables 2 and 4**). Prior to GRIN, 25 genes were common to both SA-EUR and SA-MDD (**Fig. 4**). Following GRIN, 11 genes were commonly retained from both sets of summary statistics, 8 genes were commonly removed, and 6 genes were removed or retained from one set only (**Fig. 4; Supplementary Tables 8-9**). Next, we compared ToppGene gene set enrichments before and after GRIN to investigate if GRIN functionally refined these gene sets. At *p* < 1e^-5^, 243 enrichments were significant from the unfiltered SA-EUR gene set, including 33 transcription factor binding sites and 114 GO enrichments (**Supplementary Table 10**). GRIN retained highly interrelated genes from SA-EUR as demonstrated by 274 significant enrichments, including 132 significantly enriched GO terms and 44 transcription factor binding sites (**Supplementary Table 11**). Conversely, only two enrichments were obtained from removed SA-EUR genes, indicating little functional interrelatedness among these genes (**Supplementary Table 12**). Similarly, 65 enrichments were identified from SA-MDD before GRIN while 247 enrichments were identified in the GRIN retained gene set, including an enrichment for “schizophrenia,” a mental health disorder which presents an increased risk of suicidality (DisGeNET C0036341; **Supplementary Tables 13-14**). Conversely, only 31 enrichments were identified in the removed gene set (**Supplementary Table 15**). This strongly indicated that GRIN-filtered SA-EUR and SA-MDD gene sets contained highly interrelated genes that were relevant to neurobiological pathways.

**Figure 4.**
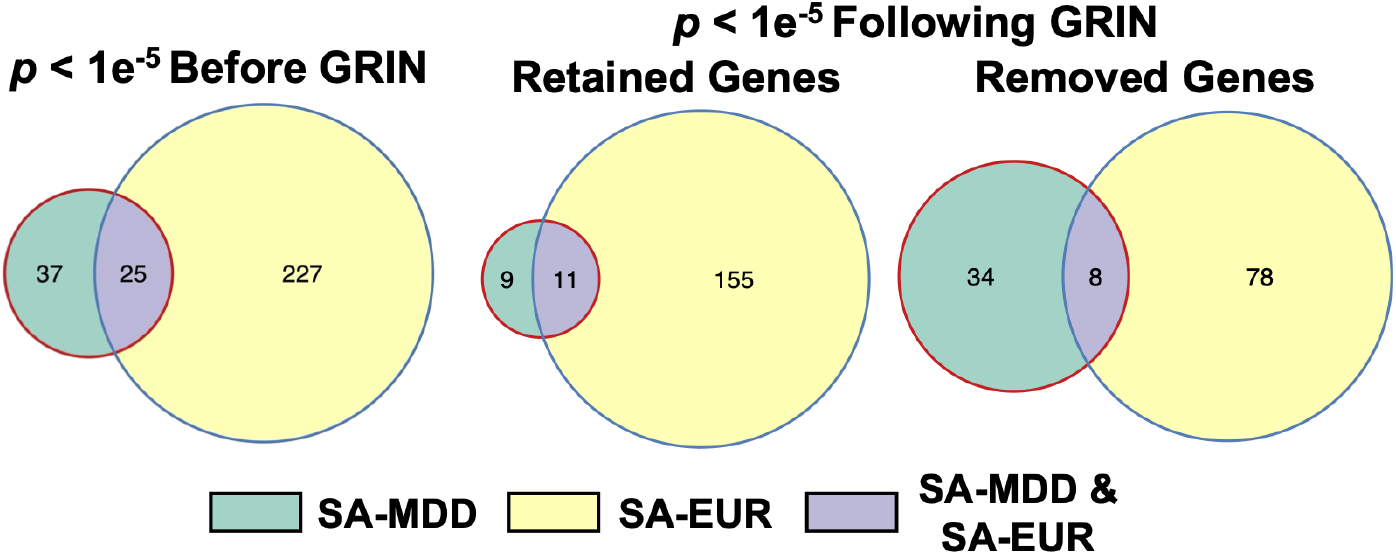
GRIN retains and removes genes common to two sets of civilian suicide attempt summary statistics. Venn diagrams of the number of genes before GRIN (left) and retained and removed genes following GRIN from suicide attempt of general European population (SA-EUR) and conditioned on major depressive disorder diagnosis (SA-MDD) at *p* < 1e^-5^. A majority of the retained genes from SA-MDD were also retained from the SA-EUR summary statistics.

### GRIN retains a majority of genes from the union of the MVP and ISGC summary statistics, enhancing functional gene set enrichment

In a process similar to a meta-analysis, we applied GRIN to the union of MVP and ISGC genes to identify replicated genes across cohorts and identify unified biological mechanisms. Prior to applying GRIN, at *p* < 1e^-5^ one gene (*PDE4B*) was common among all results, three genes were common to only MVP and SA-EUR, and 24 genes were common to only SA-EUR and SA-MDD (**Fig. 5**). After applying GRIN to the union of all genes, 17 genes common to multiple summary statistics were retained (over 50%), including *PDE4B*, 2 out of 3 common genes between MVP and SA-EUR, and 14 out of 24 common genes between SA-EUR and SA-MDD (**Fig. 5, Supplementary Table 16**). Conversely, 8 genes between SA-EUR and SA-MDD and 1 gene between MVP and SA-EUR were commonly removed at this threshold (**Fig. 5, Supplementary Table 16**).

**Figure 5.**
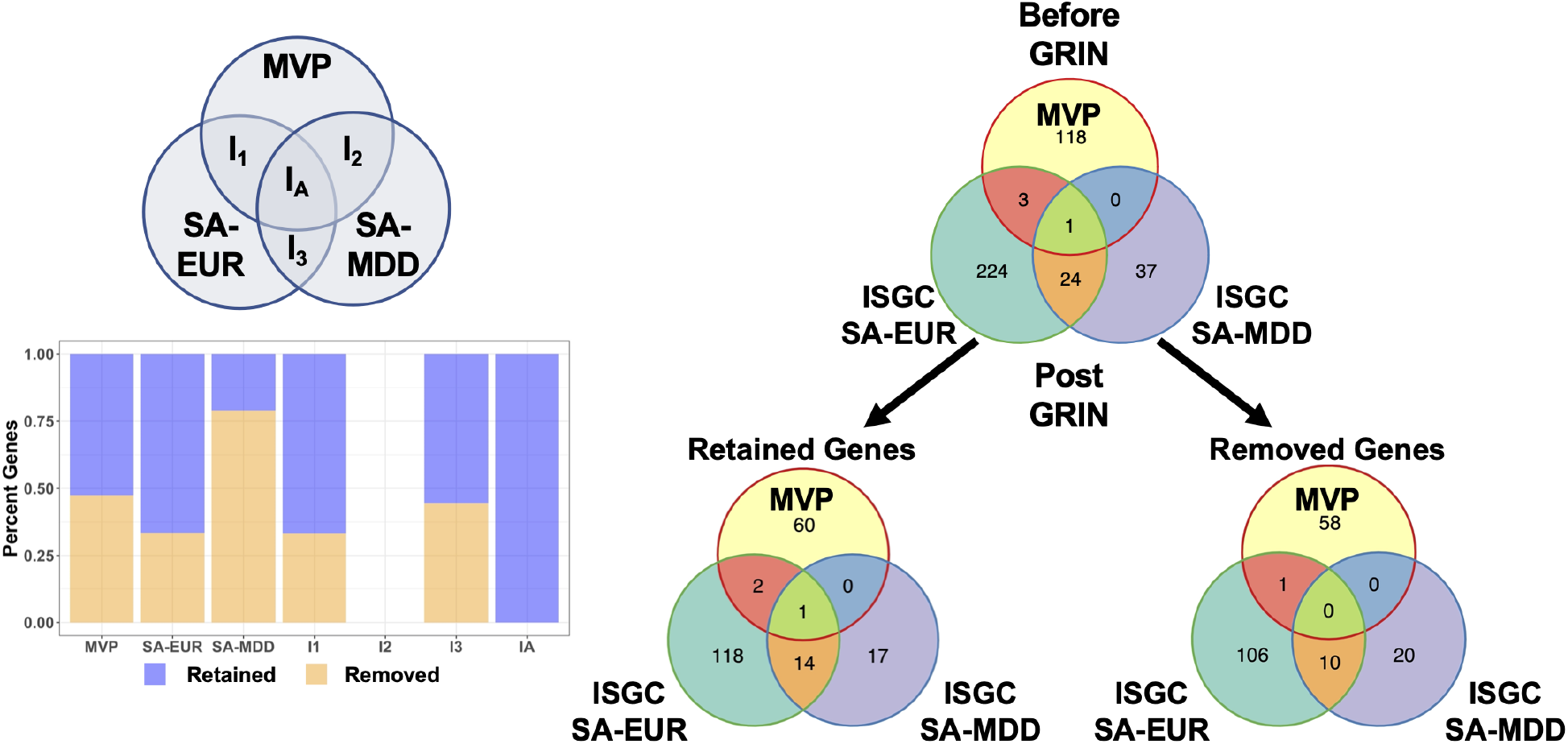
GRIN retains a majority of genes common among distinct suicide attempt summary statistics. Venn diagrams of GRIN retained and removed genes from the union of MVP, SA-EUR, and SA-MDD suicide attempt summary statistics from genes assigned from SNPs at *p* < 1e^-5^. Most genes common to multiple summary statistics were retained, as indicated by percentage of retained (orange) and removed (blue) genes in multiple sets of summary statistics. I1 = overlapping genes between MVP and SA-EUR, I2 = overlapping genes between MVP and SA-MDD, I3 = overlapping genes between SA-EUR and SA-MDD, IA = genes common to all summary statistics.

Next, we assessed GRIN’s ability to improve gene set enrichment analysis using genes common to two or more sets of summary statistics. Prior to GRIN there were 1324 significant enrichments from the 28 genes common to multiple data sets (**Fig. 6, Supplementary Tables 16-17**). Intriguingly, 1443 significant enrichments were obtained from the 17 genes commonly retained by GRIN in multiple data sets, including 126 GO terms such as “dopaminergic synapse” (**Fig. 6, Supplementary Table 18**). Conversely, only 358 enrichments were significant from the 11 intersecting genes removed by GRIN, indicating lower interrelatedness among these genes compared to the retained set (**Supplementary Table 19**). In addition to introducing newly enriched terms, GRIN improved the significance of the majority of enriched terms from the pre-filtered gene set. Of the 1324 enrichments obtained from the unfiltered set of genes common to multiple suicide attempt summary statistics, 1259 of these enrichments were retained; 1168 of these retained enrichments were more significant following GRIN, strongly indicating that retained genes constituted a more biologically cohesive set (**Fig. 6, Supplementary Tables 18-19**).

**Figure 6.**
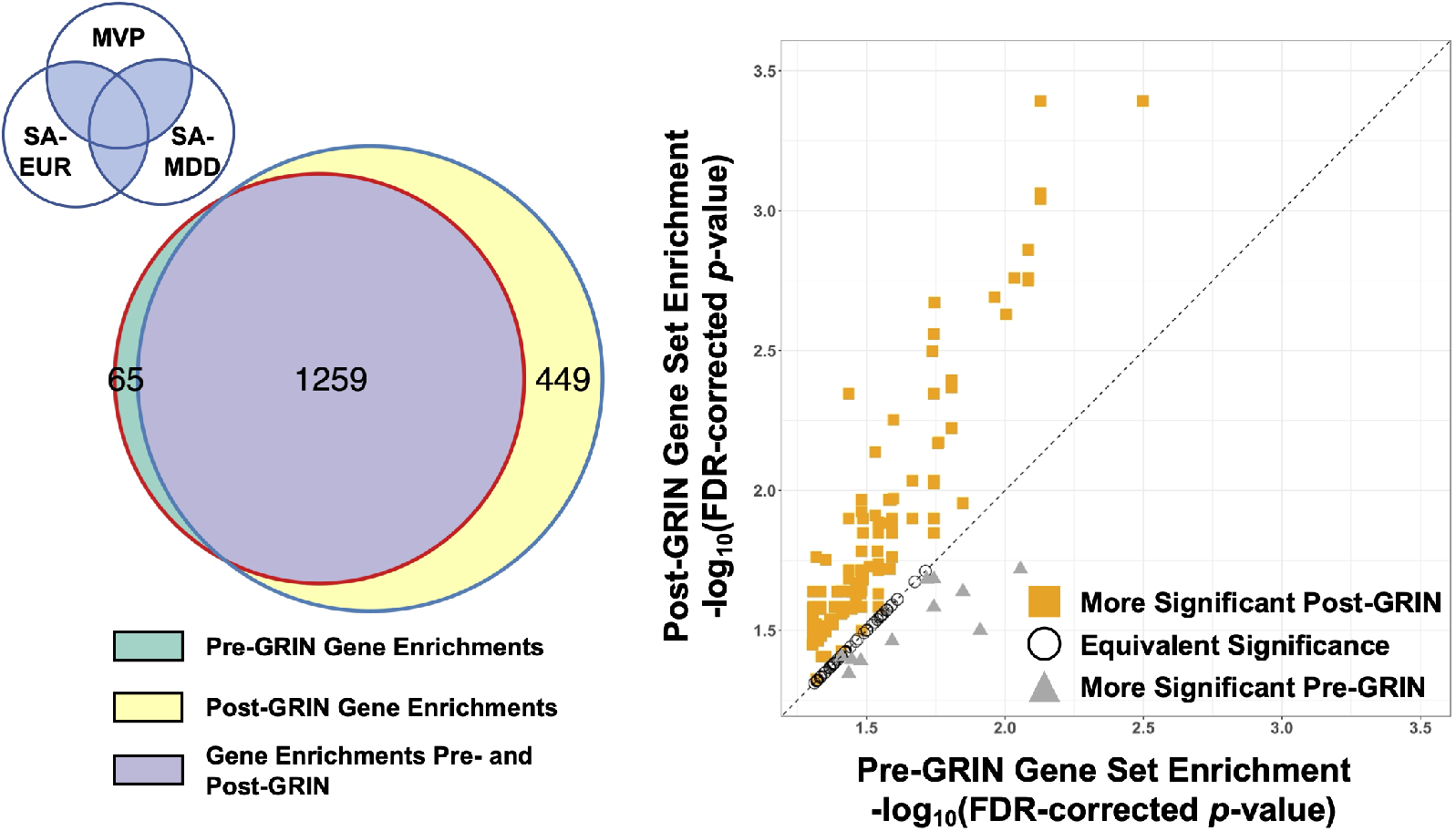
GRIN enhances gene set enrichment analysis. Left: Using genes intersecting in two or more GWAS summary statistics (shaded blue region of Venn diagram) prior to GRIN resulted in 1324 enrichments. Intersecting genes retained by GRIN resulted in 1708 enrichments, 1259 of which were common prior to GRIN. Green: Enrichments using intersecting genes prior to GRIN; yellow: enrichments using intersecting genes from GRIN retained genes; purple: enrichments common to unfiltered and GRIN-retained genes. Right: Of the 1259 enrichments common before and after applying GRIN, these enrichments were likely to be more statistically significant (1048 enrichments, orange squares) compared to equivalent significance (205 enrichments, open circles) or less significance (gray triangles). Diagonal line indicates equivalent *p*-values before and after GRIN. Threshold for enrichment inclusion was – log_10_(FDR-corrected *p*-value) > 1.30103 (FDR-corrected *p*-value < 0.05).

Similarly, when applying GRIN to each set of summary statistics separately, 13 intersecting genes were commonly retained and 9 intersecting genes were commonly removed (**Supplementary Fig. 3, Supplementary Table 20**). This resulted in more numerous and statistically significant gene set enrichments using intersecting GRIN-retained genes compared to the unfiltered gene set or GRIN-removed genes (**Supplementary Fig. 4, Supplementary Tables 21-22**).

### Retained genes from GRIN identify putative pathophysiological pathways involved in suicide attempt, leading to drug repurposing/side effect candidates

Using genes retained by GRIN, we identified biological pathways implicated by suicide attempt GWAS (**Fig. 7**). Only one of the three genes identified from MVP GWAS at *p* < 5e^-8^ (*TSHZ2*) was retained by GRIN at *p* < 1e^-5^. Multiple genes identified in both cohorts were relevant to dopaminergic signaling, including the dopamine D2 receptor subunit (*DRD2*) and phosphodiesterase 4-beta (*PDE4B*) as well as a protein kinase A subunit (*PRKAR2A*) from MVP, which can subsequently modulate cAMP/CREB-mediated transcription of genes important for synaptic plasticity^13^. Additionally, *SGIP1* was retained by GRIN in MVP and SA-EUR summary statistics, which affects presynaptic vesicle release and emotional state^14,15^. Furthermore, genes involved in neurotransmitter release (*BRSK1*) and glutamatergic synapses (*CELSR3*^*16*^) were identified along with *ICE2*, which is induced by NMDA receptor activity^17,18^. *NCAM1* was also retained by GRIN, which is a crucial mediator of synaptic plasticity and memory processes^19,20^. Multiple genes involved in cytoskeletal reorganization were also identified including *CDC42BPB, MAP4*, and *MARK3*. Moreover, *TSHZ2, SMARCC1*, and *ZNF589* were retained by GRIN and have been implicated in neurodevelopmental processes while *RCOR1* is important for neural progenitor differentiation into neuronal and glial subtypes^13,21–24^. Finally, a number of genes involved in global translation processes were identified (*DALRD3, DHX30*, and *EIF5*), two of which have been previously implicated in neurodevelopmental disorders arising from missense variants^25,26^ (**Fig. 7**).

**Figure 7.**
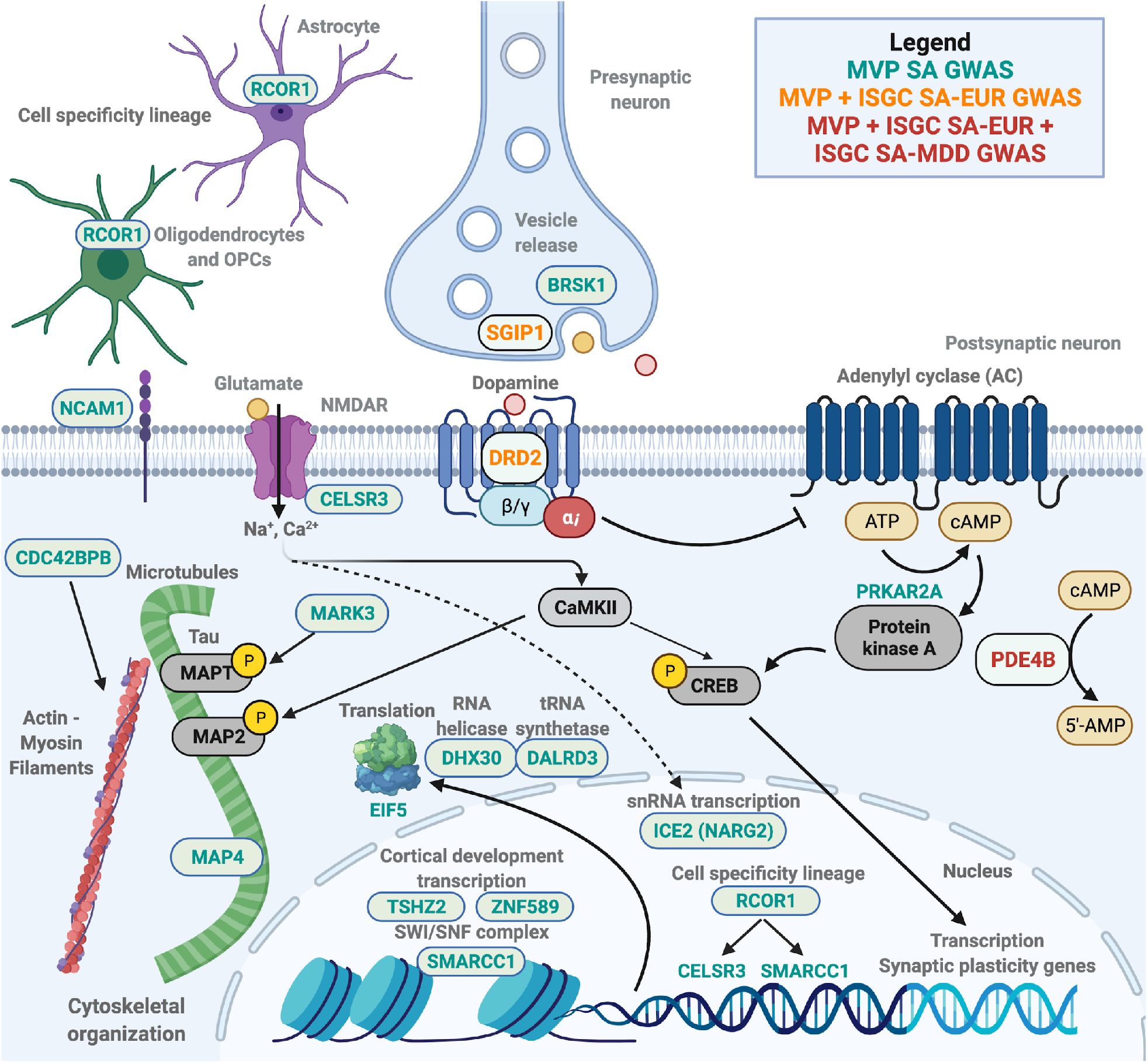
Neurobiological mechanisms implicated in suicide attempt pathophysiology as determined by GWAS summary statistics and GRIN. GRIN retained genes from MVP suicide attempt GWAS (cyan), MVP and ISGC general population GWAS (orange), and MVP and both ISGC suicide attempt summary statistics (red). A number of genes related to dopaminergic signaling (*DRD2, PRKAR2A*, and *PDE4B*) were identified, as well as presynaptic vesicle release (*BRSK1, SGIP1*), glutamatergic synapse formation (*CELSR3*), and synaptic plasticity (*NCAM1*). Genes related to cytoskeletal reorganization (*CDC42BPB, MAP4*, and *MARK3*) were also implicated in suicide attempt GWAS. Genes related to chromatin reorganization (*SMARCC1*), cortical development (*TSHZ2, ZNF589*), cell lineage (*RCOR1*), and translation processes (*DALRD3, DHX30*, and *EIF5*) were also retained by GRIN. Together, GRIN retained genes from multiple coincident biological processes underlying suicide pathophysiology. Figure made with BioRender.com.

Finally, we identified drugs that may modulate suicidal behavior based on GWAS-implicated genes retained by GRIN. Multiple drugs target the dopamine D2 receptor subunit, including the FDA-approved drugs clozapine (used to prevent suicidal behavior in schizoaffective individuals^27^) and amisulpride (**Fig. 8, Supplementary Table 23**). Roflumilast and a number of other molecular compounds also directly affect *PDE4B*. Furthermore, fostamatinib is known to affect 8 genes implicated in both MVP and SA-EUR suicide attempt GWAS. These drug-gene target links warrant future studies to ensure that they do not present increased risk for suicidality as a side effect, and to evaluate candidates for drug repurposing for suicide prevention.

**Figure 8.**
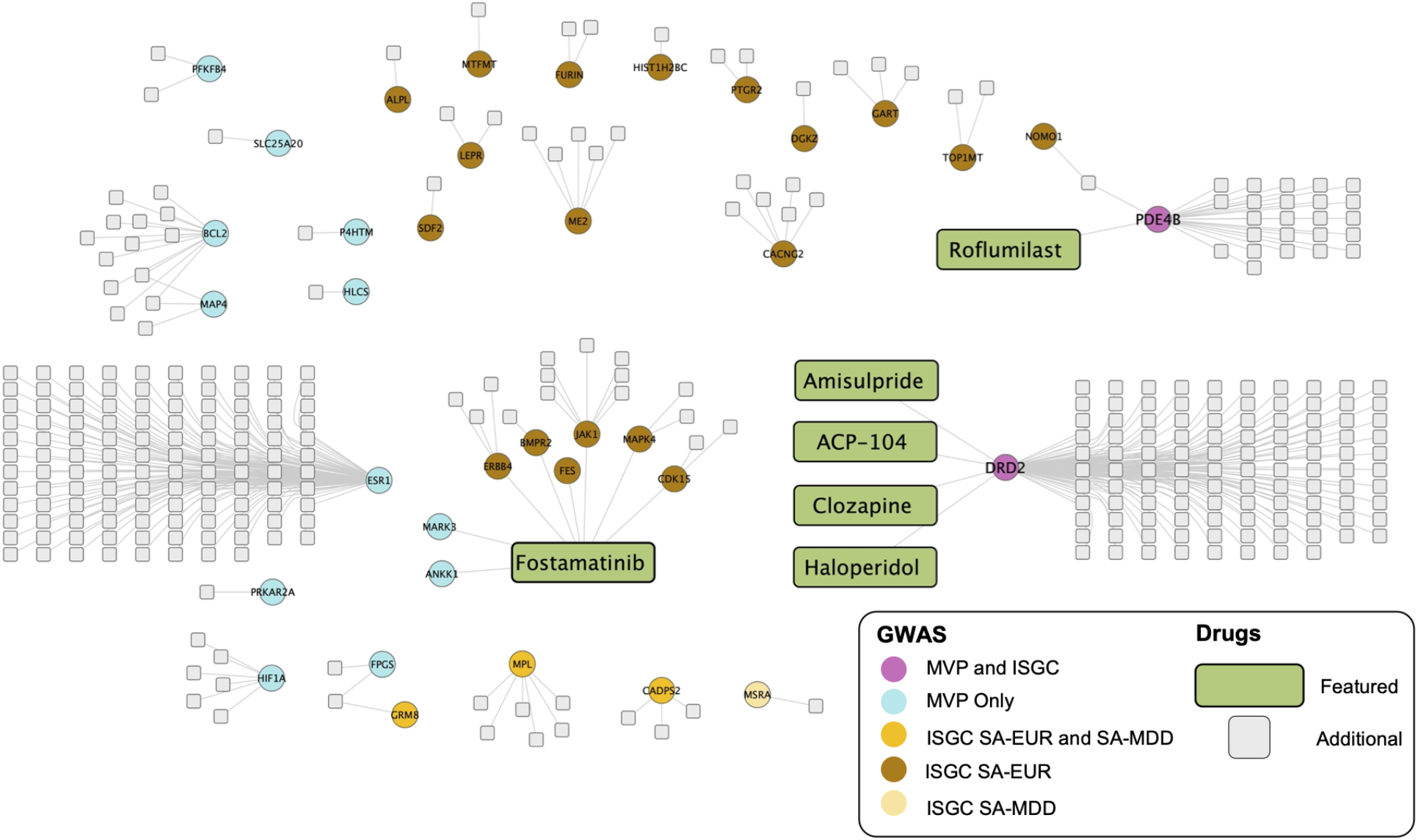
Network association of drugs and drug targets identified by suicide attempt GWAS. Network diagram of DrugBank drugs (featured drugs green, additional drugs gray) and drug target genes from genes retained by GRIN applied to MVP and ISGC suicide attempt summary statistics at *p* < 1e^-5^ threshold. The dopamine D2 receptor (*DRD2*), phosphodiesterase 4B (*PDE4B*), and estrogen receptor 1 (*ESR1*) are targeted by many drugs, while fostamatinib targets 8 genes implicated by suicide attempt GWAS followed by GRIN. Magenta: genes implicated in MVP and both SA-EUR and SA-MDD ISGC summary statistics; cyan: genes implicated in MVP only; orange: genes implicated in both SA-EUR and SA-MDD; brown: genes implicated in SA-EUR only; yellow: genes implicated in SA-MDD only.

## Discussion

Here we introduced GRIN, a workflow based on networks of biological relationships to enable the relaxing of GWAS thresholds while reducing the impact of false positives, thus producing an expanded prioritized gene set compared to a GWAS using the standard significance threshold. Genome-wide association studies are subject to statistical challenges that have historically made it difficult to identify a large proportion of trait-relevant genes. In human studies, genes that may contribute to disease have traditionally been identified from SNP associations at a genome-wide significance threshold of *p* < 5e^-8^, but for many traits GWAS often fails to find a comprehensive signal at this stringent threshold, especially complex traits controlled by many small-effect loci. Yet we know that much of the signal of heritability lies below this threshold. Relaxing the stringency gives access to more SNPs (and hence more genes) that may be associated with the trait, at the risk of introducing an increasing proportion of false positives that will confound downstream analyses. GRIN operates on the concept that genes affecting the same trait are likely to have some sort of functional relationship, even if based upon minor and/or indirect effects. Given a set of genes from a GWAS, true positive genes should therefore be functionally related to each other in some way, while false positive genes will be functionally distant with respect to each other and with respect to the true positive genes. We demonstrated that GRIN accurately partitions an input gene set into two subsets based on this mechanism and used it to boost GWAS signals. In contrast to other network-based propagation tools for enhancing GWAS (*e*.*g*., NAGA^28^, uKIN^29^, GWAB^30^), GRIN simultaneously enhances GWAS sensitivity while reducing the risk of false positive genes from being included.

The first stage of GRIN requires a representation of known relationships between all genes in a network format. Here we used a biological multiplex network which captures a wide variety of relationship types across its 10 layers. One could instead use a pre-existing network based on the species of interest, such as HumanNet^31^, YeastNet^32^, or AraNet^33^. However, GRIN’s capacity to accurately refine gene sets is limited by the connectivity represented in the network. A poor network lacking sufficient biological relationships would result in reduced ability to distinguish between functionally related genes and random noise. We therefore included various experimental data sources and generated a frontal cortex-specific predictive gene expression network using an explainable-AI methodology (iRF-LOOP^34^), which can define relationships that were not present in the literature. This novel, tissue-specific weighted network is a powerful community resource, as iRF-LOOP-derived networks can contain more informative relationships compared to traditional gene co-expression networks by an order of magnitude^34^.

Determining the functional relatedness between the genes in a GWAS gene set can be performed by many algorithms that operate on a biological network. Network propagation algorithms use information encoded in the topology of each layer simultaneously, providing a systems-level view of gene-to-gene relationships. Utilizing one such algorithm (random walk with restart, RWR), we achieved high AUROC, AUPRC, precision, recall, and specificity. Future studies are warranted to explore which sources of experimental data (*e*.*g*., protein-protein interaction, transcription factor binding) are most influential for retaining biologically interrelated genes with GRIN.

We demonstrated that GRIN works with simulated noisy gene sets similar to what is obtained from a GWAS or differential gene expression experiment. GRIN successfully partitioned curated gene sets spiked with random genes into signal and noise subsets, even when given multiple functional groups or a high noise ratio. The results confirm that RWR indeed ranks the functional group(s) of genes highly while random genes mostly receive poor rankings. When applied to real-world data, it is up to GRIN’s second stage to determine the optimal cutoff point that divides functional genes from false positive genes. The strong simulated test results provide confidence that when GRIN is applied to GWAS results, the true positive genes should rise to the top of the rankings as long as they are more functionally related to each other than random genes are, thus providing a retained set that has a higher signal-to-noise ratio than if GRIN was not used at all.

After applying GRIN to expanded GWAS results at *p* < 1e^-5^, we obtained more gene set enrichments in the retained set with consistently higher levels of significance, despite this set being considerably smaller. Thus, GRIN removed false positives that were diluting the functional signal in the original gene set. The fact that the removed gene set was scarcely enriched supports this argument. By separating a gene list into retained and removed subsets, the user can identify additional biologically relevant pathways that may be missed by enrichment analyses on the whole set alone due to dilution with noise.

While we demonstrated that GRIN achieves high accuracy using gold standard gene sets, GRIN sometimes discarded true positive genes. This indicates that not all genes removed by GRIN are necessarily irrelevant to the trait or disease, and the removed set should be considered but with lower confidence than the retained set. It is also important to consider the possibility for false positives and false negatives to be re-classified as new experimental data sources become available. For example, some genes removed by GRIN currently have few experimental gene-gene network relationships (*e*.*g*., the non-coding RNA *RP11-839D17*.*3* and pseudogene *MRPS21P1*). However, future experiments may identify their capacity to modulate transcriptional or post-transcriptional processes with pathophysiological implications. Therefore, GRIN output should be considered as guidance rather than a definitive determination of what is a true or false positive.

Combined with multiple SNP-to-gene assignments, incorporating SNPs at a less stringent significance threshold and applying GRIN elucidated additional suicide-associated genes and pathways. In addition to conventional MAGMA, we leveraged frontal cortex-specific chromatin accessibility and H-MAGMA which refined SNP-to-gene assignment. While certain variants have been previously described (*e*.*g*., variants in *DRD2, PDE4B*, and *SPATA17*^*8,9,11*^), the present study characterizes additional genes contributing to dendritic structure and multiple key neurotransmitter pathways associated with suicidality. Among these additional genes, missense variants in *CDC42BPB*^*35*^, *DALRD3*^*26*^, *DHX30*^*25*^, *SMARCC1*^*22*^, and *ZNF589*^*36*^ are known to impair behavioral and neurodevelopmental processes. While the genetic variants in the present study did not include these missense or loss-of function variants, it is possible that the variants implicated in the present suicide attempt summary statistics may alter the transcription of these genes. For example, the retained gene sets were enriched for multiple transcription factor binding sites, including an NFAT transcription factor binding site enrichment common to each GWAS based on different genes (TGGAAA, TRANSFAC NFAT_Q4_01; **Supplementary Tables 7, 11, and 14**). Variants leading to differential gene transcription may have downstream developmental or behavioral consequences leading to increased impulsivity and ultimately an increased risk of suicidality. In addition, multiple genes were implicated in cytoskeletal reorganization. *CDC42BPB* encodes MRCKbeta, a protein kinase that is induced by long-term potentiation in rodent models and mediates dendritic spinogenesis by actin-myosin filament phosphorylation^37,38^. *MAP4* is a microtubule-associated protein (MAP) and *MARK3* has been shown to phosphorylate tau (*MAPT*), another MAP which accumulates in multiple neurodegenerative disorders^39,40^. Moreover, *RCOR1* is a subunit of the REST/CoREST complex and has been shown to affect *CELSR3* and *SMARCC1* transcription in mouse models^16,22^, and *SMARCC1* has been implicated in autism as a core component of the SWI/SNF complex^23,41^. These findings point to the possible pleiotropic nature of these genes being associated with multiple neurological disorders.

By lowering the significance threshold and applying GRIN to refine suicide GWAS gene sets, we identified previously characterized drug targets (*DRD2, MARK3*, and *PDE4B*) and drug repurposing/side effect candidates that would not have been detected otherwise. Notably, the *DRD2* antagonist clozapine is the only FDA-approved drug with on-label use to prevent suicidal behavior^27^. Amisulpride is also a *DRD2* antagonist that has been shown to exhibit antipsychotic and antidepressant activities^42^. Intriguingly, fostamatinib targets 8 genes implicated by suicide attempt GWAS including *MARK3* and *PDE5B*, a different phosphodiesterase than the *PDE4B* gene implicated in suicide attempt GWAS^43^. Moreover, it is important to understand if drugs can present adverse side effects modulating suicidal behavior. For example, the *PDE4B* inhibitor roflumilast has a rare adverse side effect of increased suicidality in some individuals^44^. Further studies are warranted to understand how pharmacological manipulation of these GWAS-implicated drug targets affect the propensity of suicidal behaviors in at-risk individuals.

GRIN is a powerful tool for identifying biologically interrelated genes and for identifying true positive variants and associated genes from GWAS. In effect, GRIN synthesizes multiple lines of evidence to determine which genes should be investigated further, automating a task that researchers usually do manually post-GWAS. By applying this tool to multiple GWAS results, we identify new genes involved in suicide pathophysiology that may lead to important clinical insights.

## Data Availability

GWAS summary statistics from the Million Veteran Program used in this study will be made available on the NIH database of Genotypes and Phenotypes (dbGaP) under accession ID phs001672.v1.p. Summary statistics from the International Suicide Genetics Consortium are available at https://tinyurl.com/ISGC2021. GRIN code and the multiplex network used for this study is located at https://github.com/sullivanka/GRIN.

## Acknowledgements and Disclosures

This research used resources of the Oak Ridge Leadership Computing Facility at the Oak Ridge National Laboratory, which is supported by the Office of Science of the U.S. Department of Energy under Contract No. DE-AC05-00OR22725. GWAS summary statistics from the Million Veteran Program used in this study will be made available on the NIH database of Genotypes and Phenotypes (dbGaP) under accession ID phs001672.v1.p. Summary statistics from the International Suicide Genetics Consortium are available at https://tinyurl.com/ISGC2021. The authors declare no competing interests.

## Author Contributions

Conceptualization: KAS, DK, DAJ; Methodology: KAS, DK, DAJ; Software: KAS, ML, MCM, JIM, AC, JR, DK; Formal analysis: KAS, DK; Investigation: KAS, DK, XQ, JL, NM, DAJ; Resources: DR, XQ, DAJ; Writing -Original Draft: KAS, DK; Writing - Review & Editing: KAS, ML, MP, AMW, NM, AD, HC, DMR, MRG, AEAK, JCB, BM, DWO, NAK, DK, DAJ; Visualization: KAS, DK; Supervision: NAK, DAJ, DK; Project administration: AD, HC, DMR, JPP, AEAK, JCB, BM, DWO, NAK, DAJ; Funding acquisition: DMR, DK, DAJ.

## Online Methods

### Multiplex biological network generation

In order to capture biological relationships from diverse types of biological evidence, a multiplex network was assembled from weighted network connections (edges) from a combination of publicly available and newly generated monoplex (single layer) networks. A multiplex network has an advantage over aggregate multilayer networks in that the unique topology of each layer is maintained, resulting in generally higher functional predictive ability^45^. Multiple component networks from HumanNet v2^31^ were used (co-functional links by co-citation, co-essentiality^46^, co-expression, molecular pathway databases, gene neighborhood, phylogenetic profile associations, and orthologous protein-protein interactions transferred from model organisms [CC, CE, CX, DB, GN, PG, IL]), and a protein-protein interaction (PPI) network was generated by merging the following networks into a single monoplex layer: HumanNet v2 component PPI networks (HT, LC), and high-confidence physical protein-protein interactions from STRING version 11.0^47^ (taxa = 9606, protein.actions.v11.0, mode=binding, min score = 700). A transcription factor-gene network layer was included based on a previously published, human brain-specific transcription factor binding site network^48^. Newly generated Predictive Expression Networks (PENs) were obtained using the Iterative Random Forest - Leave One Out Prediction (iRF-LOOP) method^34,49^ using individual-level RNA-seq expression data from the Genotype-Tissue Expression (GTEx) project^50^ from the frontal cortex (Brodmann area 9). The resulting multiplex network was built using RWRtoolkit (https://github.com/dkainer/RWRtoolkit), which incorporates command-line scripts and an R library for generating multiplex networks and running the network exploration algorithm random walk with restart (RWR) by building upon the RandomWalkRestartMH R package^45^. The multiplex network used for all analyses comprises 10 layers, 51,183 unique genes, and 3,419,975 edges using δ = 0.5, where δ is the probability of the random walker remaining in the current network layer or moving to a different layer. The multiplex network used for all analyses is publicly available at https://github.com/sullivanka/GRIN/tree/main/test/suicide_weighted_Multiplex_0.5Delta.RData.

### GRIN process

GRIN leverages the hypothesis that false positive genes in gene sets are likely to be functionally random with respect to the rest of the gene set, while true positive genes are likely to share function with other members of the gene set. Using this theory, GRIN partitions a gene set, such as from differential gene expression analysis or SNP-to-gene assignment from GWAS, into retained and removed genes in a two-stage process.

In Stage 1, every gene in the network is ranked according to how connected it is to the genes in the user-specified gene set (*e*.*g*., GWAS-derived genes). This includes ranking the user-specified genes themselves by using leave-one-out cross-validation (LOOCV). GRIN can make use of any network exploration algorithm that can provide gene rankings. We chose RWR for its robustness, but other propagation methods (*e*.*g*., PageRank, heat diffusion) can also achieve this^28,29,51^. RWR provides each gene with a rank that is a proxy for how easily each gene in the network can be reached from the starting set of GWAS genes, including a rank for the GWAS genes themselves. Genes with many paths and interactions to one or more of the GWAS genes rank strongly, while genes that are isolated or distant from the GWAS genes rank poorly. In the current implementation, this RWR-based ranking occurs based on network propagation of probabilities of visiting a given gene in the multiplex network, which is based on a matrix representation of the edge weights between genes in the multiplex network (i.e., the supra-adjacency matrix composed of all intra-and inter-layer connections). Random walks are then simulated many times by propagating the probability of the random walker exploring a given gene beginning from the seed genes, and this process continues until the combined network probabilities no longer change between simulated random walks by a given threshold (1e^-10^), thereby achieving convergence based upon an asymptotic number of simulated random walks. The advantage of using a propagation algorithm like RWR is that genes that are not direct neighbors of GWAS genes may still rank highly due to indirect paths. Additional parameters can be used to tune RWR to favor certain network layers (τ) or adjust the probability of restart (*r*) at seed genes. In all analyses in the present study, we used *r* = 0.7, equivalent τ values for all network layers, and a multiplex network with δ = 0.5 based on previous work that achieved good performance using these parameters^45^.

To obtain accurate rankings for each gene in a gene set of size *n*, we chose to implement random walk with restart leave-one-out cross validation (RWR-LOOCV) *n* times, where in each run one gene is left out and the other *n-1* genes are used as seed genes (starting points) for the random walker in the multiplex network. Each run of RWR-LOOCV generates a ranking of every non-seed gene in the multiplex, including the left-out gene from the original seed gene set, so that each gene in the user’s obtains *n-1* rank values after *n* runs. Stage 1 then orders the genes in the set from best to worst according to their median rank values. GRIN also needs a representation of what Stage 1 results should look like for purely random gene sets of size *n*. This empirical null distribution is generated by running RWR-LOO for 100 gene sets, each containing *n* randomly sampled genes from the multiplex. The median rank at each position in the order from 1 to *n* thus represents the empirical null distribution of ranks for this specific multiplex and gene set size.

In Stage 2, a cutoff *C* between 1 and *n* is determined below which all gene set members are considered the equivalent of random and can be discarded. A two-sided Mann-Whitney U test from the R stats base package (“wilcox.test”) is performed over a sliding window of size *winsize* = 0.15 × *n* to see if the RWR-LOOCV ranks for the gene set members come from the same distribution as the null distribution RWR-LOOCV ranks. The expectation is that a gene set window containing functional groups of genes will have a very different ranking distribution to the random genes in the equivalent null window, resulting in very small (significant) *p*-values. On the other hand, if the window contains genes with little functional relatedness, the ranking distribution will appear to be drawn from the null distribution and the *p*-value tends towards 1.

This test is run for each window sliding by 1, producing a *p*-value vector of length *n*-*winsize*. The cutoff *C* is chosen by finding an elbow in the *p*-values using the open source R package “Knee Arrower” with the method = “first” parameter set (https://github.com/agentlans/KneeArrower). The output is a *retained* gene set and a *removed* gene set.

### Validation of GRIN using well-characterized gene sets

To determine the ability of GRIN to effectively remove noise genes from a gene set, we obtained a variety of well-characterized (“gold”) biologically interrelated gene sets and spiked them with random genes drawn from the full multiplex network. Given our application of this method to suicide GWAS summary statistics, we chose 20 gene sets related to diverse brain functions. We included an additional 10 gene sets related to other organ systems (lung and kidney) in order to demonstrate that GRIN could be adapted to other biological contexts. These thirty “gold standard” gene sets of functionally interrelated genes (see **Supplementary Table 1**), ranging in size from 10 to 225 genes, were derived from the following sources: Gene Ontology (GO^52^); Online Mendelian Inheritance in Man (OMIM^53^); and DisGENET^54^. Random genes were inserted into each gold set to create gene sets with a 1:1 signal-to-noise ratio (i.e. N_gold_ : N_random_). For each of the 30 gold sets we generated 100 test gene sets using varying samples of random genes. GRIN was then used to filter out random genes from each test gene set and the effectiveness of the filter was evaluated using receiver operator characteristics (ROC) and precision/recall (PR) measured at every possible cutoff point, *C*, in each rank-ordered gene set.

For evaluation purposes, “true positive” genes were labeled as genes belonging to a gold gene set that were correctly retained by GRIN; “true negative” genes were randomly added genes that were correctly removed by GRIN; “false positive” genes were randomly added genes that were incorrectly retained by GRIN; and “false negative” genes were gold genes that were incorrectly removed by GRIN. ROC (false positive rate vs true positive rate), and PR curves (precision vs recall) were generated and area under ROC (AUROC) and area under PRC (AUPRC) values were calculated for each test gene set. Median AUROC and AUPRC were calculated for each of the 30 gold standard gene sets to indicate whether Stage 1 of GRIN ranked gold genes more highly than random genes in general. After estimating the optimal cutoff *C* at Stage 2, precision, recall, and specificity (true negatives / true negatives + false positives) were calculated for the genes removed and the genes retained by Stage 2. Median precision, recall, and specificity values were calculated across the 100 test gene sets for each of the 30 gold standard gene sets. Values are presented as median +/- interquartile range (IQR).

GRIN was also tested on unequal ratios of gold standard genes and random genes using the dopaminergic synaptic signaling gene set from GO (GO:0001963) and Acute Kidney Failure gene set from DisGeNET (C0022660) – 2:1 gold genes to random genes, 1:2 gold genes to random genes, and 1:4 gold genes to random genes. For each ratio of gold standard genes to noise, 100 test sets were generated. Notably as precision increases (reduction of false positives), there is a compensatory decrease in recall, as there is the possibility of throwing out too many true positives, leading to an increased number of false negatives. Since there is a higher precision value at lower noise levels, there is an inverse trend of decreasing recall at lower noise levels.

Finally, to test whether GRIN could remove random genes from gene sets containing multiple groups belonging to biological processes that were functionally distinct, multiple gold gene sets were combined and random noise also added. Dopaminergic synaptic transmission (GO:0001963; 23 genes) and central nervous system myelination (GO:0022010; 20 genes) were mixed with two ratios of random to gold standard genes – 2:1 gold genes:noise and 1:1 gold genes:noise. This process was repeated to generate 100 gene sets of gold standard and random genes for each ratio examined.

### Million Veteran Program (MVP) suicide attempt genome wide association study (GWAS) summary statistics

Suicide attempts were identified from United States veterans as described previously^11^.

Suicide attempts were characterized by using a combination of Veterans Healthcare Administration (VHA) databases from the VA: the Suicide Prevention Application Network (SPAN) database, electronic health record (EHR) information from the VA Corporate Data Warehouse (CDW), and the CDW Mental Health Domain survey. For the MVP diagnosis, suicide attempt was determined by the presence of one or more of the following International Statistical Classification of Diseases and Related Health Problems (ICD)-9 and ICD-10 diagnostic codes in a subject’s EHR: ICD-9: E950-959; ICD-10: T14.91, X60-62, X64, X66-X83, Y87.0, Z91.5. Control patients were obtained from veterans enrolled in MVP without a history of suicide attempt or suicidal ideation as determined by a combination of SPAN survey, Mental Health Domain survey, and ICD diagnostic codes in the CDW database (suicidal ideation codes: ICD-9: V62.84; ICD-10: R45.851). A total of 410,464 controls from various ancestries (African, Asian, European, and Hispanic) were included for genome-wide association along with 14,535 cases of non-fatal suicide attempt and 294 fatal attempts. Genome-wide association analyses were conducted using DNA from whole blood samples from subjects enrolled in MVP using a custom Affymetrix Biobank Array. Quality control and imputation was performed as previously described^10^. All subjects provided informed consent and the activities used to generate the GWAS summary statistics were approved by the VA Central Institutional Review Board and all activities were approved by the Oak Ridge National Laboratory Institutional Review Board.

### International Suicide Genetics Consortium (ISGC) suicide attempt GWAS summary statistics

Suicide attempt summary statistics were analyzed from two sets of suicide attempt summary statistics derived from civilian populations compiled by the ISGC^9^. SNPs were included from a general population of European ancestry (SA-EUR) as 26,590 as cases of suicide attempt and 492,022 control subjects. Furthermore, additional summary statistics were derived from this general population conditioned on diagnosis status for major depressive disorder (SA-MDD) to generate an additional set of suicide attempt summary statistics. Thus, while the SA-MDD summary statistics are not independent of the SA-EUR summary statistics as they are comprised of the same set of controls and cases of suicide attempt, both SA-EUR and SA-MDD summary statistics were analyzed in order to determine the overlap between these results and results from the MVP cohort. All subjects involved in the ISGC provided informed consent and the activities used to generate the GWAS summary statistics were approved by local institutional review boards as previously described^9^ and by the Oak Ridge National Laboratory Institutional Review Board.

### SNP to gene assignment

SNPs were assigned to genes using two separate methods. H-MAGMA^1^ was used in combination with publicly available Hi-C data from adult dorsolateral prefrontal cortex (dlPFC)^55^ to improve intergenic SNP-to-gene assignment based on three-dimensional chromatin structure in this brain region. Adult prefrontal cortex Hi-C data was used as this brain region is known to be involved in executive function and impulsivity processes, which are disrupted in individuals with a history of suicide attempt^56,57^Additionally, conventional MAGMA^4^ was used as an alternate method of SNP-to-gene assignment. Thus, H-MAGMA and conventional MAGMA were applied only as methods of assigning SNPs to genes only using SNPs at given significance thresholds, rather than using these tools as gene-based tests on the entire set of summary statistics.

SNPs were assigned to genes from MVP, SA-EUR, or SA-MDD summary statistics at multiple thresholds (*p* < 5e^-8^, *p* < 1e^-5^, *p* < 1e^-4^, *p* < 1e^-3^, *p* < 1e^-2^, and *p* < 1e^-1^; **Supplementary Table 2**). The union of conventional MAGMA and H-MAGMA-assigned genes (*i*.*e*., all genes assigned from either method) from MVP, SA-EUR, or SA-MDD suicide attempt summary statistics were subsequently used as gene set inputs to GRIN at a threshold of *p* < 1e^-5^ and were filtered into retained and removed gene sets (**Supplementary Tables 5, 8, and 9**).

### Gene set enrichment analysis

Gene sets from MVP and ISGC summary statistics were tested for multiple enrichments using the online ToppGene suite using ToppFun^12^. Gene set enrichments were analyzed using the following enrichment categories: GO: Molecular Function; GO: Biological Process; GO: Cellular Component; Human Phenotype; Pathway (all databases selected); Transcription Factor Binding Site (all databases selected); Drug (all databases selected); Disease (all databases selected). Enrichments were considered significant using a Benjamini-Hochberg false discovery rate (FDR)-adjusted *p*-value threshold < 0.05.

### Drug to gene target networks for putative drug repurposing and side effect evaluation

Genes identified as contributing to suicide attempt pathophysiology from MVP and ISGC summary statistics were used to construct drug to gene target networks from information derived from DrugBank^58^. Drug to gene target networks were visualized in Cytoscape^59^ (version 3.8.2, Cytoscape Consortium) to identify drugs known to target genes of interest from MVP and ISGC summary statistics using GRIN-retained genes at *p* < 1e^-5^ (**Supplementary Table 20**). ISGC GWAS genes were compared to genes from the MVP cohort using Venn diagrams generated from the open source R package Vennerable (https://github.com/js229/Vennerable).

### Software

GRIN is available as an open-source, command-line R script for public use. The code, installation instructions, and user manual can be found at https://github.com/sullivanka/GRIN.

## References

1. Sey, N. Y. A. et al. A computational tool (H-MAGMA) for improved prediction of brain-disorder risk genes by incorporating brain chromatin interaction profiles. Nat. Neurosci. 23, 583–593 (2020).

2. Hall, M. A. et al. Novel EDGE encoding method enhances ability to identify genetic interactions. PLoS Genet. 17, e1009534 (2021).

3. Petersen, A., Alvarez, C., DeClaire, S. & Tintle, N. L. Assessing methods for assigning SNPs to genes in gene-based tests of association using common variants. PLoS One 8, e62161 (2013).

4. de Leeuw, C. A., Mooij, J. M., Heskes, T. & Posthuma, D. MAGMA: generalized gene-set analysis of GWAS data. PLoS Comput. Biol. 11, e1004219 (2015).

5. Boyle, E. A., Li, Y. I. & Pritchard, J. K. An Expanded View of Complex Traits: From Polygenic to Omnigenic. Cell 169, 1177–1186 (2017).

6. Gosak, M. et al. Network science of biological systems at different scales: A review. Phys. Life Rev. 24, 118–135 (2018).

7. Voracek, M. & Loibl, L. M. Genetics of suicide: a systematic review of twin studies. Wien. Klin. Wochenschr. 119, 463–475 (2007).

8. Erlangsen, A. et al. Genetics of suicide attempts in individuals with and without mental disorders: a population-based genome-wide association study. Mol. Psychiatry 25, 2410– 2421 (2020).

9. Mullins, N. et al. Dissecting the Shared Genetic Architecture of Suicide Attempt, Psychiatric Disorders, and Known Risk Factors. Biol. Psychiatry (2021) doi:10.1016/j.biopsych.2021.05.029.

10. Kimbrel, N. A. et al. A genome-wide association study of suicide attempts and suicidal ideation in U.S. military veterans. Psychiatry Res. 269, 64–69 (2018).

11. Kimbrel, N. A. et al. A genome-wide association study of suicide attempts in the million veterans program identifies evidence of pan-ancestry and ancestry-specific risk loci. Mol. Psychiatry (2022) doi:10.1038/s41380-022-01472-3.

12. Chen, J., Bardes, E. E., Aronow, B. J. & Jegga, A. G. ToppGene Suite for gene list enrichment analysis and candidate gene prioritization. Nucleic Acids Res. 37, W305–11 (2009).

13. Wang, H., Xu, J., Lazarovici, P., Quirion, R. & Zheng, W. cAMP Response Element-Binding Protein (CREB): A Possible Signaling Molecule Link in the Pathophysiology of Schizophrenia. Front. Mol. Neurosci. 11, 255 (2018).

14. Dvorakova, M. et al. SGIP1 is involved in regulation of emotionality, mood, and nociception and modulates in vivo signalling of cannabinoid CB1 receptors. Br. J. Pharmacol. 178, 1588–1604 (2021).

15. Trevaskis, J. et al. Src homology 3-domain growth factor receptor-bound 2-like (endophilin) interacting protein 1, a novel neuronal protein that regulates energy balance. Endocrinology 146, 3757–3764 (2005).

16. Thakar, S. et al. Evidence for opposing roles of Celsr3 and Vangl2 in glutamatergic synapse formation. Proc. Natl. Acad. Sci. U. S. A. 114, E610–E618 (2017).

17. Sugiura, N., Patel, R. G. & Corriveau, R. A. N-methyl-D-aspartate receptors regulate a group of transiently expressed genes in the developing brain. J. Biol. Chem. 276, 14257– 14263 (2001).

18. Takahashi, H. et al. MED26 regulates the transcription of snRNA genes through the recruitment of little elongation complex. Nat. Commun. 6, 5941 (2015).

19. Vukojevic, V. et al. Evolutionary conserved role of neural cell adhesion molecule-1 in memory. Transl. Psychiatry 10, 217 (2020).

20. Walmod, P. S., Kolkova, K., Berezin, V. & Bock, E. Zippers make signals: NCAM-mediated molecular interactions and signal transduction. Neurochem. Res. 29, 2015–2035 (2004).

21. Monaghan, C. E. et al. REST corepressors RCOR1 and RCOR2 and the repressor INSM1 regulate the proliferation-differentiation balance in the developing brain. Proc. Natl. Acad. Sci. U. S. A. 114, E406–E415 (2017).

22. Abrajano, J. J. et al. Differential deployment of REST and CoREST promotes glial subtype specification and oligodendrocyte lineage maturation. PLoS One 4, e7665 (2009).

23. Sokpor, G., Xie, Y., Rosenbusch, J. & Tuoc, T. Chromatin Remodeling BAF (SWI/SNF) Complexes in Neural Development and Disorders. Front. Mol. Neurosci. 10, 243 (2017).

24. Caubit, X., Tiveron, M.-C., Cremer, H. & Fasano, L. Expression patterns of the three Teashirt-related genes define specific boundaries in the developing and postnatal mouse forebrain. J. Comp. Neurol. 486, 76–88 (2005).

25. Lessel, D. et al. De Novo Missense Mutations in DHX30 Impair Global Translation and Cause a Neurodevelopmental Disorder. Am. J. Hum. Genet. 101, 716–724 (2017).

26. Lentini, J. M., Alsaif, H. S., Faqeih, E., Alkuraya, F. S. & Fu, D. DALRD3 encodes a protein mutated in epileptic encephalopathy that targets arginine tRNAs for 3-methylcytosine modification. Nat. Commun. 11, 2510 (2020).

27. Meltzer, H. Y. et al. Clozapine treatment for suicidality in schizophrenia: International Suicide Prevention Trial (InterSePT). Arch. Gen. Psychiatry 60, 82–91 (2003).

28. Carlin, D. E. et al. A Fast and Flexible Framework for Network-Assisted Genomic Association. iScience 16, 155–161 (2019).

29. Hristov, B. H., Chazelle, B. & Singh, M. uKIN Combines New and Prior Information with Guided Network Propagation to Accurately Identify Disease Genes. Cell Syst 10, 470–479.e3 (2020).

30. Shim, J. E. et al. GWAB: a web server for the network-based boosting of human genome-wide association data. Nucleic Acids Res. 45, W154–W161 (2017).

31. Hwang, S. et al. HumanNet v2: Human gene networks for disease research. Nucleic Acids Res. 47, D573–D580 (2019).

32. Kim, H. et al. YeastNet v3: a public database of data-specific and integrated functional gene networks for Saccharomyces cerevisiae. Nucleic Acids Res. 42, D731–6 (2014).

33. Lee, T. et al. AraNet v2: an improved database of co-functional gene networks for the study of Arabidopsis thaliana and 27 other nonmodel plant species. Nucleic Acids Res. 43, D996– 1002 (2015).

34. Cliff, A. et al. A High-Performance Computing Implementation of Iterative Random Forest for the Creation of Predictive Expression Networks. Genes 10, (2019).

35. Chilton, I. et al. De novo heterozygous missense and loss-of-function variants in CDC42BPB are associated with a neurodevelopmental phenotype. Am. J. Med. Genet. A 182, 962–973 (2020).

36. Agha, Z. et al. Exome sequencing identifies three novel candidate genes implicated in intellectual disability. PLoS One 9, e112687 (2014).

37. Wang, X.-X. et al. MRCKβ links Dasm1 to actin rearrangements to promote dendrite development. J. Biol. Chem. 296, 100730 (2021).

38. Li, L. et al. Protein kinases paralleling late-phase LTP formation in dorsal hippocampus in the rat. Neurochem. Int. 76, 50–58 (2014).

39. Lund, H. et al. MARK4 and MARK3 associate with early tau phosphorylation in Alzheimer’s disease granulovacuolar degeneration bodies. Acta Neuropathol Commun 2, 22 (2014).

40. Doki, C. et al. Microtubule elongation along actin filaments induced by microtubule-associated protein 4 contributes to the formation of cellular protrusions. J. Biochem. 168, 295–303 (2020).

41. Neale, B. M. et al. Patterns and rates of exonic de novo mutations in autism spectrum disorders. Nature 485, 242–245 (2012).

42. Rehni, A. K., Singh, T. G. & Chand, P. Amisulpride-induced seizurogenic effect: a potential role of opioid receptor-linked transduction systems. Basic Clin. Pharmacol. Toxicol. 108, 310–317 (2011).

43. Rolf, M. G. et al. In vitro pharmacological profiling of R406 identifies molecular targets underlying the clinical effects of fostamatinib. Pharmacol Res Perspect 3, e00175 (2015).

44. Pinner, N. A., Hamilton, L. A. & Hughes, A. Roflumilast: a phosphodiesterase-4 inhibitor for the treatment of severe chronic obstructive pulmonary disease. Clin. Ther. 34, 56–66 (2012).

45. Valdeolivas, A. et al. Random walk with restart on multiplex and heterogeneous biological networks. Bioinformatics 35, 497–505 (2019).

46. Wang, T. et al. Gene Essentiality Profiling Reveals Gene Networks and Synthetic Lethal Interactions with Oncogenic Ras. Cell 168, 890–903.e15 (2017).

47. Szklarczyk, D. et al. STRING v11: protein–protein association networks with increased coverage, supporting functional discovery in genome-wide experimental datasets. Nucleic Acids Res. 47, D607–D613 (2018).

48. Pearl, J. R. et al. Genome-Scale Transcriptional Regulatory Network Models of Psychiatric and Neurodegenerative Disorders. Cell Syst 8, 122–135.e7 (2019).

49. Basu, S., Kumbier, K., Brown, J. B. & Yu, B. Iterative random forests to discover predictive and stable high-order interactions. Proc. Natl. Acad. Sci. U. S. A. 115, 1943–1948 (2018).

50. GTEx Consortium. The Genotype-Tissue Expression (GTEx) project. Nat. Genet. 45, 580– 585 (2013).

51. Di Nanni, N., Bersanelli, M., Milanesi, L. & Mosca, E. Network Diffusion Promotes the Integrative Analysis of Multiple Omics. Front. Genet. 11, 106 (2020).

52. Harris, M. A. et al. The Gene Ontology (GO) database and informatics resource. Nucleic Acids Res. 32, D258–61 (2004).

53. Amberger, J. S., Bocchini, C. A., Schiettecatte, F., Scott, A. F. & Hamosh, A. OMIM.org: Online Mendelian Inheritance in Man (OMIM®), an online catalog of human genes and genetic disorders. Nucleic Acids Res. 43, D789–98 (2015).

54. Piñero, J. et al. DisGeNET: a comprehensive platform integrating information on human disease-associated genes and variants. Nucleic Acids Res. 45, D833–D839 (2017).

55. Wang, D. et al. Comprehensive functional genomic resource and integrative model for the human brain. Science 362, (2018).

56. Zhang, H. et al. Aberrant White Matter Microstructure in Depressed Patients with Suicidality. J. Magn. Reson. Imaging (2021) doi:10.1002/jmri.27927.

57. Cao, J. et al. The Association Between Resting State Functional Connectivity and the Trait of Impulsivity and Suicidal Ideation in Young Depressed Patients With Suicide Attempts. Front. Psychiatry 12, 567976 (2021).

58. Wishart, D. S. et al. DrugBank: a knowledgebase for drugs, drug actions and drug targets. Nucleic Acids Res. 36, D901–6 (2008).

59. Shannon, P. et al. Cytoscape: a software environment for integrated models of biomolecular interaction networks. Genome Res. 13, 2498–2504 (2003).

